# Patterns and predictors of domestic violence and abuse enquiry in South East London maternity settings: Cross-sectional analysis of routine electronic health record data collected between 2019 and 2023

**DOI:** 10.64898/2026.05.18.26353528

**Authors:** Demelza Smeeth, Roxanne C. Keynejad, Raquel Catalao, Gemma Luck, David Wood, Claire A Wilson

## Abstract

**BACKGROUND:** The UK National Institute for Health and Care Excellence recommends routine enquiry about domestic violence and abuse (DVA) in maternity care. We aimed to explore patterns and predictors of DVA enquiry during routine first antenatal care (‘booking’) appointments with midwives in South East London.

**METHODS:** We conducted an observational cohort study using cross-sectional data collected through the St Thomas’ Hospital midwifery service between 1^st^ January 2019 and 31^st^ March 2023. Pseudonymised data were extracted from maternity records, comprising demographics, mental and physical health information, social factors, and DVA enquiry. We used linear mixed modelling to test associations between predictors and DVA enquiry.

**RESULTS:** The dataset comprised 7,932 booking appointments with 7,007 women (median age: 32 years; ethnicity: 52% White, 27% Black, 7% Asian, and 15% other). Enquiry was made about current experiences of DVA in 79.4% of appointments. Black-identifying women (OR=1.28, 95% CI [1.11,1.46]) and those born in Sub-Saharan Africa (OR=1.37 [1.14,1.64]) were more likely to be asked than white-identifying and UK-born women. Single women were more likely to be asked than married or cohabiting women (OR=1.22 [1.08,1.38]). Those living in more deprived neighbourhoods were more likely to be asked (OR=1.07 [1.01,1.14]). Multivariable modelling found that being born in Sub-Saharan Africa or Southern Europe, and living alone but with additional support were all associated with increased DVA enquiry, while being born in North America or requiring an interpreter were associated with decreased enquiry

**CONCLUSIONS:** Despite recommendations for routine DVA enquiry during all booking appointments, a substantial proportion of pregnant individuals were not asked between 2019 to 2023. Predictors of DVA enquiry reflected practical barriers (e.g. language), and known or perceived predictors of DVA risk (e.g. deprivation). Our findings suggest that midwives consciously or unconsciously prioritise DVA enquiry for women they believe are at greatest risk, against national guidelines.

## Introduction

Domestic violence and abuse (DVA) is defined by the United Kingdom (UK) Home Office as abusive behaviour between people aged 16 or over, who are personally connected to one another, regardless of gender or sexuality.^1^ DVA comprises, but is not limited to, psychological, physical, sexual, financial and emotional abuse. The Crime Survey for England and Wales revealed that DVA was present in 15.4% of all crimes recorded by the police in England and Wales in the year leading up to March 2025 and 25.8% of adults (∼12.5 million people) had experienced DVA in their lifetime.^2^ DVA can begin or escalate during the perinatal period from conception until one year postpartum, increasing the risk of adverse pregnancy outcomes and maternal and neonatal death.^3,4^

However, official statistics are likely to underestimate the occurrence of DVA. Survivors of DVA face a range of barriers to disclosure, including concerns about their own or their family’s safety, feelings of self-blame and stigma.^5,6^ Disclosure and help seeking are also influenced by the isolating nature of DVA and limited opportunities to seek help. Healthcare providers play a key role in enquiring about DVA and connecting survivors to advocacy and support services.^7^ Routine healthcare appointments during pregnancy can offer such an opportunity, particularly if they are conducted in safe, confidential settings conducive to disclosure.^8^

Anyone can experience DVA regardless of age, ethnicity, socio-economic status or health status. Therefore, routine and universal enquiry is the recommended strategy to identify those experiencing DVA by the National Institute for Health and Care Excellence (NICE).^9^ However, barriers to enquiry in maternity settings include limited time, low staff confidence and lack of private space.^10^ Additional barriers might also include conscious or unconscious bias of who is more likely to be experiencing DVA.^11,12^ Limited evidence from Australia and the USA indicates that certain groups of people are more or less likely to be asked. In Australian, postpartum women were less likely to be asked if they had immigrated to Australia or were of low socioeconomic status ^13^. Conversely, in the USA, pregnant women were less likely to be asked if they were White, college-educated, or married ^14^. Concerningly DVA enquiry was also less frequent for women who had experienced DVA in the previous year.

During the COVID-19 pandemic, several indicators suggested an increased incidence of DVA in the UK and beyond.^8,15^ Police reports of DVA increased in London during the pandemic, particularly during lockdowns, with the greatest increases among women and birthing people, and people of Asian, Arab, and Middle Eastern ethnicity.^16^ However, this increase was not always reflected in healthcare settings, which may have been impacted by changes to DVA enquiry rates or practices.^17^ Maternity services underwent several changes to the ways in which care was delivered during the COVID-19 pandemic, with fewer consultations, more virtual appointments and restrictions on the presence of accompanying partners.^18^ During this time, midwives expressed concerns about the safety of discussing sensitive topics during remote consultations, especially via telephone.^19,20^ However, others pointed out benefits to DVA enquiry of pregnant individuals attending appointments alone.^19^ Whether these changes impacted DVA enquiry is currently unknown.

We aimed to explore patterns and predictors of DVA enquiry during routine maternity booking appointments held between January 2019 and March 2023in South East London. First, we sought to describe the rate of DVA enquiry during these appointments. Second, we investigated how enquiry rates changed over time, with particular attention to the period of the COVID-19 pandemic. Based on the evidence of reduced DVA identification in healthcare settings during the pandemic,^17^ we hypothesised that DVA enquiry would decline during this period. Finally, we aimed to identify predictors of DVA enquiry in maternity care. We hypothesised that DVA enquiry would be less likely among women facing greater structural or interpersonal barriers to disclosure, including younger women, women from minoritised ethnic backgrounds, and those living in more deprived areas. This expectation reflects known inequities in healthcare interactions and the potential influence of bias on which women midwives choose to ask about DVA.

## Methods

### Study design and data source

This was an observational cohort study using cross-sectional data collected through the antenatal care service provided via St Thomas’ Hospital in the London borough of Lambeth between 1^st^ January 2019 and 31^st^ March 2023. Lambeth is an ethnically diverse inner city borough with high levels of deprivation and a larger proportion of younger adults compared with averages in London and England.^21,22^

Pseudonymised data were extracted from electronic maternity records on the BadgerNet system, created during routine booking appointments for individuals who registered their pregnancy with St Thomas’ Hospital. The booking appointment is the first appointment with the midwifery service during a pregnancy in the UK and usually occurs before 12 weeks’ gestation. Data collected during the booking appointment included demographic information (age, ethnicity), information about mental and physical health and risk factors for adverse pregnancy outcomes, including current and previous experiences of DVA victimisation. Data were accessed by the research team for research purposes in 20/05/2025.

This research was conducted to support and contextualise the activities of Lambeth Early Action Partnership (LEAP).^23^ LEAP was one of five local partnerships that formed a ten-year (2015-2025) programme which aimed to improve the life chances of babies, young children, and families in some of Lambeth’s most economically deprived areas. A formal data sharing agreement (DSA) between LEAP and Guy’s and St Thomas’ NHS Foundation Trust supported the sharing of pseudonymised maternity booking data with LEAP for service monitoring and research and evaluation on the legal basis of legitimate interest. Data storage was confidential and adhered to the DSA and Kings College London Research Data Management Policy standards. The King’s College London Psychiatry, Nursing, and Midwifery Research Ethics Sub-committee approved the study (LRS/DP-23/24-42972).

### Exposures

We reviewed variables collected during the booking appointment for their inclusion as potential predictors of DVA enquiry in this study. We selected variables based on the literature and plausibility. We also included variables which we hypothesised would act as practical barriers to DVA enquiry. We did not select variables if they were largely missing (>25%), were not well-defined, overlapped considerably with another variable, were recorded inconsistently, or were not reasonably hypothesised to impact DVA enquiry.

We defined the pandemic as the period from 24^th^ March 2020 to 18^th^ April 2022. The dates were chosen to encompass the period of time from the first government-legislated national lockdown until social distancing was no longer required in National Health Service (NHS) hospitals and general practice (GP) surgeries. We defined lockdown periods by stay at home orders present in periods of national lockdown and tier 4 restrictions when social distancing was legally mandated.^24^

### Demographic variables

Ethnicity was recorded as Office for National Statistics recommended categories (White, Black, Asian, mixed and other). Country of birth was self-reported and converted to United-Nations-defined regions using *whoville* and *countrycode* R packages.^25,26^ The primary language spoken at home was self-reported and used as a binary variable (English vs any other language) to account for those who spoke English as a second language. Whether an interpreter was required was recorded. Age was calculated from date of birth and the booking date. We defined adolescent pregnancy as women aged under 18 years and advanced maternal age as women aged 35 years or older at the booking appointment

### Socioeconomic variables

Pregnant women self-reported whether they were entitled to public funds (most commonly British or Irish citizens, refugees, and those with humanitarian protection or Settled Status). rea deprivation was defined using the Townsend deprivation index. The Townsend deprivation index is an area-based measure of material deprivation based on unemployment, overcrowding, non-car ownership, and non-home ownership derived from the person’s ward of residence using 2011 census data from the UK data service.^27^ Employment status was reported as one of five categories: employed, caring responsibilities, no right to work, student and unemployed. Pregnant women self-reported their current accommodation. We used this variable as a five-level variable: owner-resident, private rental, housing association or council rental, living with friends of family, and no fixed abode. Pregnant women also self-reported whether they had any problems with their housing.

### Social variables

Pregnant women self-reported their current marital status. We used this as a categorical variable with three levels: married or cohabiting; single; or widowed, divorced or separated. Pregnant women reported their current support status with regards to who they were living with. This was used as a categorical variable with six levels: living with partner or family; living with other family or friends; living alone, but with additional support; living alone, unsupported; in-patient facility or in care; or in sheltered accommodation. Pregnant women self-reported whether they feel currently supported and whether they had any current relationship issues.

### Reproductive and health variables

Pregnant women were asked two brief questions (Whooley questions) to detect potential depression and used as binary variables.^29^ Women also reported if they had seen a psychiatrist before or if their partner had any mental health problems. Pregnant women reported if they were current smokers and if there were any smokers in their household. Pregnant women self-reported alcohol consumption between conception and booking. Finally, women were flagged by midwives for medical risk factors if they had any chronic health problem which could impact the pregnancy including diabetes, hypertension or obesity. Women self-reported whether they had previously been pregnant and whether the current pregnancy was planned. We defined a late booking as having a booking appointment later than 20 weeks’ gestation.^28^

### Outcome: DVA enquiry

During booking appointments, pregnant individuals were asked whether they were currently experiencing DVA, with responses recorded as yes, no, declined to answer, or unable to ask. For the purposes of this study, DVA enquiry was dichotomised into enquiry made (responses of yes, no, or declined to answer) or no enquiry made (when midwives reported being unable to ask).

### Statistical analyses

All analyses were conducted in R (v. 4.4) in RStudio.^30^ To address item-level missingness, the dataset was imputed using multiple imputation by chained equations (MICE) with the *mice* package using 20 iterations and 20 chains.^31^ This resulted in 20 complete datasets, with missing values filled in with plausible estimates derived from observed data distributions. All analysis variables were included in the imputation models, in addition to the following auxiliary variables: alcohol consumption before conception, enquiry regarding past experiences of DVA, and English-speaking ability. We selected imputation methods which were recommended for each variable type. We visually assessed convergence of imputed values across the 20 iterations through trace-plots and compared imputed values with observed data using diagnostic plots to ensure validity. We applied analyses to all 20 imputed datasets to take into account the inherent uncertainty produced by imputation and pooled the results using Rubin’s rules.^32^

We used linear mixed modelling to test associations between each predictor and DVA enquiry using binomial generalized linear mixed models fitted with the *lme4* package ^33^. All continuous variables were scaled and centred for analysis. We analysed temporal trends and those associated with the pandemic by modelling time as either a continuous (years) or categorical variable (pre-pandemic, pandemic and post-pandemic), accounting for midwifery team as a random effect to account for team-specific differences in DVA enquiry. We explored non-linear trends in DVA enquiry over time by implementing cubic spline modelling using the *splines* package which is a data-driven approach to model temporal data defined piece-wise by polynomials.^34^ We adjusted for midwifery team as a random effect and allowed eight degrees of freedom which best fitted the data. Using the resulting model, we predicted DVA enquiry over time for plotting.

We explored potential predictors of DVA enquiry using binomial generalized linear mixed models. Predictors were tested in separate models, with booking date as a fixed effect and midwifery team as a random effect.

Many of the predictors investigated were strongly related to one another (e.g. ethnicity and region of birth) which precluded direct multivariable modelling. Therefore, we performed feature selection to select variables for multivariable modelling using elastic-net penalized regression implemented in the *glmnet* R package ^35^. This approach shrinks the contribution of less important variables toward zero, selecting the most relevant variables and preventing overfitting when many predictors are considered at once. Numerical variables were standardised and dummy variables were created for categorical variables prior to analysis. This process was repeated over each of the 20 imputed datasets. Variables with non-zero coefficients across all imputed datasets were considered selected features to be used in multivariable modelling. The final multivariable model was created using binomial generalized linear mixed models as described previously.

## Results

### Cohort description

Between 1^st^ January 2019 and 31^st^ March 2023, 7,932 booking appointments for 7,007 different women were held across NHS maternity services covered by St Thomas’ Hospital. A cohort description can be found in Table 1. Booking appointment attendees had a median age of 32 years (IQR: 29,36). Reflecting the multicultural setting, the cohort was 51.5% White, 26.5% Black, 7.2% Asian, 8.1% mixed, and 6.7% other ethnicities. The median gestation at booking was 69 days (IQR: 60, 85) and 65.2% of the cohort had been pregnant at least once before.

**Table 1.** Cohort description.

|  |  |
| --- | --- |
| N booking appointments | 7932 |
| Enquiry about current DVA, n (%) | 6299 (79.4) |
| Age at booking, median (IQR) | 32 (29, 36) |
| Adolescent pregnancy, n (%) | 42 (0.5) |
| Ethnicity, n (%) |  |
| White | 4082 (51.5) |
| Black | 2105 (26.5) |
| Asian | 573 (7.2) |
| Mixed | 640 (8.1) |
| Other | 531 (6.7) |
| Born in the UK, n (%) | 3846 (48.5) |
| Interpreter required, n (%) | 593 (7.5) |
| Townsend deprivation index, median (IQR) | 9.0 (7.4, 9.6) |
| Marital status, n (%) |  |
| Married or cohabiting | 4951 (62.4) |
| Single | 2923 (36.9) |
| Widowed or divorced | 58 (0.7) |
| Pregnant before, n (%) | 5173 (65.2) |
| Planned pregnancy, n (%) | 5341 (67.3) |
| Gestation at booking (days), median (IQR) | 69 (60, 85) |
| Late booking, n (%) | 620 (7.8) |
| Smoker at booking, n (%) | 294 (3.7) |
| Drank alcohol since conception, n (%) | 215 (2.7) |

Based on imputed dataset.

### Trends in DVA enquiry

In 79.4% of booking appointments an enquiry about current DVA was made, while in 20.6% of appointments midwives reported being unable to ask. The rate of enquiry about current DVA gradually decreased over the study period (OR=0.94/year, 95% CI: 0.90,0.98), although large fluctuations were apparent from cubic spline fitted modelling (Figure 1). The COVID-19 pandemic covered 48% of the study period. The pandemic period was associated with a reduction in enquiry (pre-pandemic vs pandemic: OR=0.87, 95% CI: 0.77,0.99), which fell further during the post-pandemic period (pre-pandemic vs post-pandemic: OR=0.70, 95% CI: 0.59,0.81). However, following the end of 2022 the rate of DVA enquiry increased to 81.9%, suggesting a return to pre-pandemic DVA enquiry levels (Figure 1).

**Figure 1.**
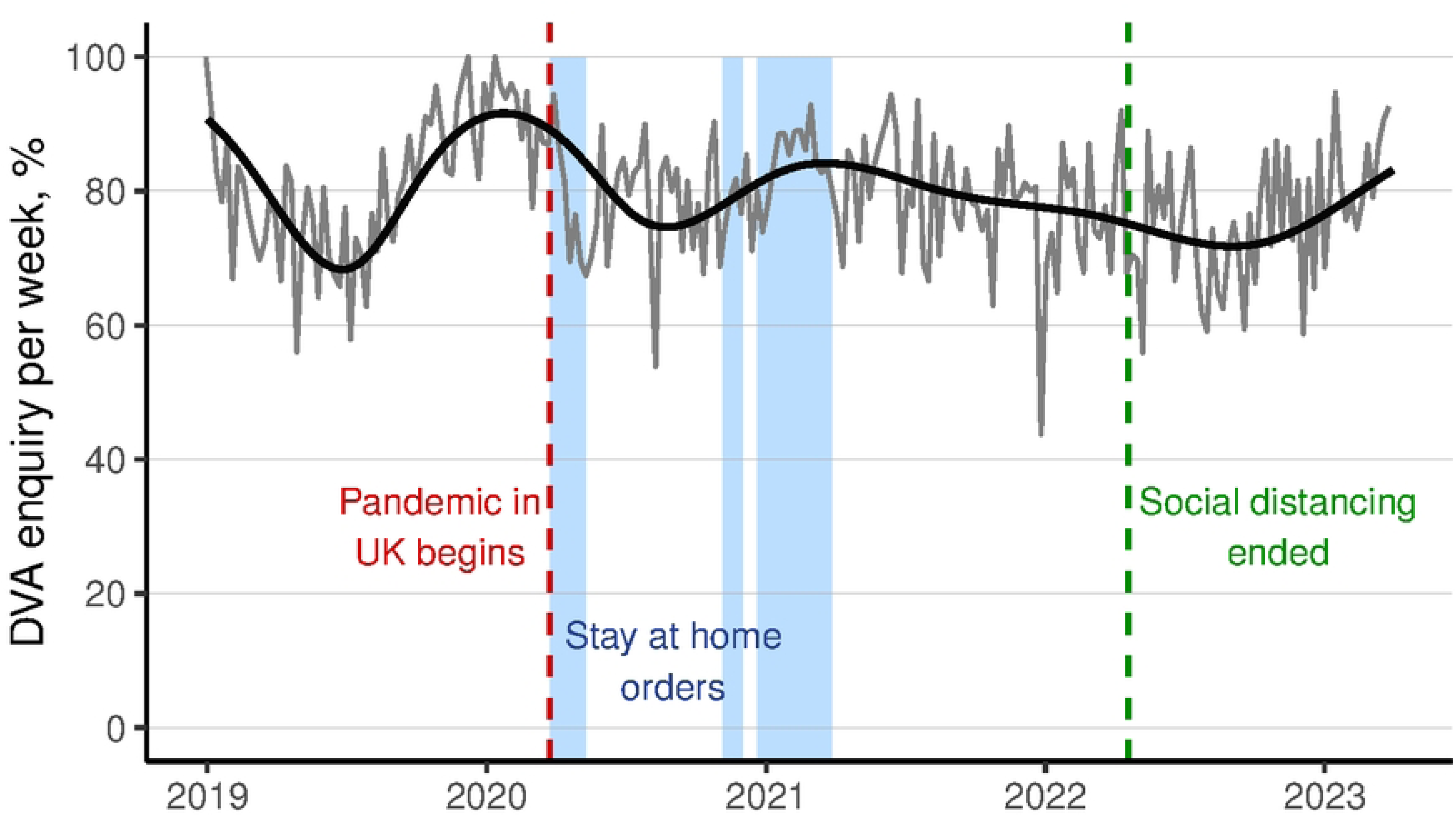
Rate of DVA enquiry per week. All datapoints in grey represent data collected from at least 10 booking appointments. Smoothed predicted probability of DVA enquiry over date based on a spline model in black. The beginning and end of the COVID-19 pandemic marked in periods of national lockdown and tier 4 restrictions when social distancing was legally mandated marked in blue.

### Demographic predictors of DVA enquiry

Several demographic factors were associated with the likelihood of midwives enquiring about current experiences of DVA during booking appointments (Table 2). Compared to those who self-identified as White, Black-identifying individuals were more likely to be asked about current DVA victimisation (OR=1.28, 95% CI: 1.11,1.46, p<0.001). Similarly, those who were born in Sub-Saharan Africa were more likely to be asked than those born in the UK (OR=1.37, 95% CI: 1.14,1.64, p<0.001). Those who were born in Southern Europe were also more likely to be asked about current DVA than UK-born individuals (OR=1.34, 95% CI: 1.07,1.68, p=0.016), while those who were born in North America were less likely to be asked (OR=0.59, 95% CI: 0.42,0.83, p=0.002). While maternal age was not linearly associated with DVA enquiry, women who were under the age of 18 years (adolescent pregnancies) at their booking appointment were less likely to be asked about DVA (OR=0.50, 95% CI: 0.25,1.00, p=0.049).

**Table 2.** Demographic predictors of enquiry about current DVA.

|  | OR [95% CI] | p |
| --- | --- | --- |
| Age at booking<sup>a</sup> | 1.00 [0.99,1.01] | 0.818 |
| Adolescent pregnancy<sup>b</sup> | 0.50 [0.25,1.00] | 0.049 |
| Advanced maternal age<sup>b</sup> | 0.99 [0.88,1.11] | 0.793 |
| Ethnicity (ref. White) |  |  |
| Black | 1.28 [1.11,1.47] | <0.001 |
| Asian | 1.04 [0.83,1.29] | 0.758 |
| Mixed | 0.96 [0.78,1.18] | 0.714 |
| Other | 1.01 [0.80,1.27] | 0.924 |
| Region of birth (ref. UK) |  |  |
| Australia and New Zealand | 1.24 [0.80,1.93] | 0.342 |
| Central Asia | 3.70 [0.48,28.4] | 0.208 |
| Eastern Asia | 0.68 [0.45,1.04] | 0.073 |
| Eastern Europe | 1.09 [0.82,1.45] | 0.574 |
| Latin America and the Caribbean | 1.10 [0.90,1.35] | 0.330 |
| Northern Africa | 1.11 [0.65,1.90] | 0.696 |
| Northern America | 0.59 [0.42,0.83] | 0.002 |
| Northern Europe | 1.12 [0.75,1.67] | 0.576 |
| South-Eastern Asia | 1.07 [0.65,1.76] | 0.790 |
| Southern Asia | 1.45 [0.97,2.15] | 0.067 |
| Southern Europe | 1.34 [1.07,1.68] | 0.016 |
| Sub-Saharan Africa | 1.37 [1.14,1.64] | <0.001 |
| Western Asia | 1.26 [0.80,1.83] | 0.321 |
| Western Europe (exc. UK) | 1.32 [0.95,1.83] | 0.098 |
| Primary language at home not English<sup>b</sup> | 1.08 [0.95,1.22] | 0.226 |
| Interpreter required<sup>b</sup> | 0.84 [0.69,1.03] | 0.093 |

All models adjusted for booking date as a fixed effect and midwifery team as a random effect. <sup>a</sup>Continuous variables scaled and centred. <sup>b</sup>Binary variables reference group: no.

### Socioeconomic predictors of DVA enquiry

Some socioeconomic indicators were associated with the likelihood of midwives enquiring about current experiences of DVA (Table 3). Neighbourhood deprivation (Townsend deprivation index) was associated with the likelihood of DVA enquiry with those living in more deprived neighbourhoods more likely to be asked (OR=1.07, 95% CI: 1.01,1.14, p=0.024). Women who were living in rented accommodation (private or local authority) or had no fixed abode were more likely to be asked about their current experiences of DVA than those who own their home (OR=1.23 95% CI: 1.07,1.42, p=0.004; OR=1.35, 95% CI: 1.17,1.57, p=6.7×10^−5^; OR=1.61, 95% CI: 1.11,2.34, p=0.012). However, having access to public funds, employment status and housing problems were not robustly associated with current DVA enquiry.

**Table 3.** Socioeconomic predictors of enquiry about current DVA.

|  | OR [95% CI] | p |
| --- | --- | --- |
| Townsend deprivation index<sup>a</sup> | 1.07 [1.01,1.14] | 0.024 |
| Access to public funds<sup>b</sup> | 0.86 [0.53,1.38] | 0.529 |
| Employment (ref. employed) |  |  |
| Caring responsibilities | 1.06 [0.83,1.35] | 0.656 |
| No right to work | 1.48 [0.50,4.35] | 0.450 |
| Student | 0.94 [0.65,1.38] | 0.728 |
| Unemployed | 1.16 [0.99,1.35] | 0.065 |
| Accommodation (ref. owner-resident) |  |  |
| Private rental | 1.23 [1.07,1.42] | 0.004 |
| Housing association or council rental | 1.35 [1.17,1.57] | <0.001 |
| With friends or family | 1.17 [0.95,1.44] | 0.150 |
| No fixed abode | 1.60 [1.09,2.35] | 0.012 |
| Housing problems<sup>b</sup> | 1.18 [0.92,1.50] | 0.193 |
| All models adjusted for booking date as a fixed effect and midwifery team as a random effect. |  |  |
| <sup>a</sup>Continuous variables scaled and centred. <sup>b</sup>Binary variables reference group: no. |  |  |

### Social predictors of DVA enquiry

Social factors were associated with DVA enquiry (Table 4). Compared to those who were married or cohabiting with a partner, single women were more likely to be asked about current DVA victimisation (OR=1.22, 95% CI: 1.08,1.38, p=0.002). Compared to those who were living with their partner or family, those who were living alone but still reported additional social support were more likely to be asked (OR=1.61, 95% CI: 1.27,2.04, p<0.001). However, feeling supported and reports of relationship issues were not associated with likelihood of DVA enquiry.

**Table 4.** Social predictors of enquiry about current DVA.

|  | OR [95% CI] | p |
| --- | --- | --- |
| Marital status (ref. married or cohabiting) |  |  |
| Single | 1.22 [1.08,1.38] | 0.002 |
| Widowed, divorced or separated | 1.85 [0.79,4.36] | 0.157 |
| Support status (ref. lives with partner or family) |  |  |
| Lives with other family or friends | 1.22 [0.87,1.72] | 0.247 |
| Lives alone, supported | 1.61 [1.27,2.04] | <0.001 |
| Lives alone, unsupported | 1.27 [0.81,1.99] | 0.294 |
| In-patient/In-care | 0.19 [0.02,2.08] | 0.174 |
| Sheltered accommodation | 2.04 [0.87,4.82] | 0.104 |
| Feeling supported<sup>a</sup> | 0.66 [0.42,1.04] | 0.071 |
| Relationship issues<sup>a</sup> | 1.60 [0.92,2.80] | 0.097 |
| All models adjusted for booking date as a fixed effect and midwifery team as a random effect. |  |  |
| <sup>a</sup>Binary variables reference group: no. |  |  |

### Reproductive and health predictors of DVA enquiry

Some reproductive factors were also associated with DVA enquiry (Table 5). Those who had been pregnant before were more likely to be asked about current DVA than women attending for their first pregnancy (OR=1.18, 95% CI: 1.05,1.32, p=0.005). Conversely, women who reported having planned their pregnancy were less likely to be asked about current DVA (OR=0.78, 95% CI: 0.68,0.89, p=2.4×10^−4^). Having a late booking appointment (>20 weeks of gestation) and advanced maternal age (>35 years old) were not associated with DVA enquiry.

**Table 5.** Reproductive and health predictors of enquiry about current DVA.

|  | OR [95% CI] | p |
| --- | --- | --- |
| Pregnant before | 1.18 [1.05,1.32] | 0.005 |
| Planned pregnancy | 0.78 [0.68,0.89] | <0.001 |
| Late booking | 0.88 [0.73,1.08] | 0.223 |
| PHQ: Feeling down, depressed or hopeless | 1.00 [0.83,1.19] | 0.977 |
| PHQ: Little interest or pleasure in doing things | 0.98 [0.80,1.21] | 0.848 |
| Seen a psychiatrist before | 0.88 [0.69,1.14] | 0.345 |
| Partner mental health problems | 0.77 [0.61,0.96] | 0.021 |
| Smoker at booking | 1.31 [0.96,1.79] | 0.091 |
| Smokers present in household | 1.12 [0.94,1.34] | 0.204 |
| Any alcohol after conception | 1.01 [0.67,1.54] | 0.952 |
| Medical risk factors | 1.09 [0.98,1.23] | 0.126 |
| All models adjusted for booking date as a fixed effect and midwifery team as a random effect. All variables reference group: no. |  |  |

Finally, very few health-related variables were associated with enquiry about current DVA (Table 5). Only those who reported having a partner with mental health problems were less likely to be asked about current DVA (OR=0.77, 95% CI: 0.61,0.96, p=0.021). Current symptoms of depression, having seen a psychiatrist, current smoking status, smokers in the home, recent alcohol use and medical risk factors were not associated with DVA enquiry.

### Multivariable models

Elastic net regression selected 48.5% (15/31 variables) of the tested variables for multivariable modelling (Supplementary Table 2). In multivariable models, ethnicity was no longer associated with DVA enquiry but women who were born in Sub-Saharan Africa or Southern Europe were still more likely to be asked about current DVA after adjustment for other factors (adjusted OR=1.25, 95% CI: 1.01,1.55, p=0.039; aOR=1.36, 95% CI: 1.07,1.72, p=0.016). Conversely, women born in North America were less likely to asked about their experiences of DVA in multivariable models (aOR=0.65, 95% CI: 0.46,0.91, p=0.013). Women who required an interpreter in the appointment were also less likely to be asked, after adjusting for other factors (aOR=0.73, 95% CI: 0.58,0.91, p=0.007), while women who live alone but with additional support were more likely to be asked (aOR=1.38, 95% CI: 1.07,1.78, p=0.014). All other variables selected for multivariable modelling were not significantly associated with DVA enquiry after adjusting for other factors.

## Discussion

Despite recommendations for routine DVA enquiry during all booking appointments where practical, a substantial proportion of pregnant individuals were not asked about it at their booking between appointment 2019 and 2023. Booking appointment data revealed that the rate of enquiry across the full time period of the cohort was 79.4%, but this fell to as low as 40 to 60% during some weeks. There was also evidence that the rate of enquiry has fallen gradually over time, but we were unable to differentiate a general trend from the specific impact of the COVID-19 pandemic due to significant fluctuations in the rate of DVA enquiry. From early to mid-2020, social distancing was recommended, many booking appointments were held virtually, and the NHS faced staff shortages with concomitant impacts on the workload and morale of existing staff.^19^ These alterations to clinical service provision could feasibly have impacted DVA enquiry. However, we observe that enquiry returned to pre-pandemic levels during 2023. Due to data availability, it was not possible to monitor beyond early 2023, but more recent data would help to determine whether DVA enquiry has truly returned to pre-pandemic levels.

DVA was not asked about uniformly across different groups, and several demographic, socioeconomic, social and reproductive factors were associated with the likelihood of being asked. Some predictors of reduced DVA enquiry may reflect practical barriers facing midwives. The fact that women who required an interpreter for their appointment were less likely to be asked may arise from general communication problems or confidentiality concerns in the presence of a third party. Language barriers are an established obstacle to seeking help for DVA and also represent a key mechanism by which individuals may be further isolated by their abuser.^6^ This finding is particularly important given that migrant women with language barriers have been highlighted by the confidential inquiries into maternal deaths in the UK, indicating additional obstetric and psychiatric risks and care barriers.^36^ Black, minoritised and migrant women are also recognised as being at elevated risk of gender-based violence^37^, as well as experiencing intersectional barriers to DVA disclosure.^38^ This finding therefore highlights the potential for system improvements and training interventions to optimise care for this group of women, in addition to research exploring barriers to enquiry.

Women who were single were more likely to be asked about their experiences of DVA than those who were married or cohabiting, similar to US findings.^14^ Women in a relationship may be more likely to be accompanied by their partner to their booking appointment, and midwives have previously identified this as a barrier to enquiry.^10^ Together, these findings emphasise the importance of standardising procedures for routine enquiry in the absence of partners, for example during weighing outside the consultation room.

Many predictors of increased DVA enquiry found in this cohort and in the USA^14^ were established predictors of DVA risk itself, for example, unplanned pregnancy or living in an area of deprivation. Women with an unplanned pregnancy are almost three times more likely to experience DVA compared with women with a planned pregnancy,^39^ while reports of DVA are higher among women living in more versus less deprived areas.^40^ Some factors, such as teenage pregnancies and unplanned conceptions may frequently co-occur, intensifying this risk.^41^ Other predictors may not be confirmed risk factors for DVA but may be assumed to be such. These findings suggest that midwives may consciously or unconsciously prioritise DVA enquiry for women who they judge to be at elevated risk, despite universal enquiry being the recommendation and findings that indicate that women who are at greater risk of DVA may be less likely to be asked.^14^ Qualitative research is needed to explore this hypothesis with clinical staff and interrelationships with other factors such as staff workload, burnout and own experiences of DVA.^42^

This study has several strengths. As far as we know, the is the first investigation of its kind in Europe. By using routinely collected electronic health record data we were able to comprehensively examine associations between DVA enquiry and a range of demographic, social and economic factors. We also avoided reliance on clinicians’ self-reports of DVA enquiry or their perceived reasons for non-enquiry. While midwives may report barriers to DVA enquiry, many of these may not be apparent to practitioners or may not be reported due to deviating from national guidelines. Electronic health records also enabled data from over 6000 appointments to be analysed, allowing consideration of rarer predictors, such as adolescent pregnancy.

This study also had some limitations. Firstly, we could only examine patterns of DVA enquiry for a four-year period. This prevented us from identifying longer term trends in enquiry and how demographic changes in the pregnant population may influence the relationships observed. Secondly, the data are limited to booking appointments. The antenatal care pathway offers multiple opportunities for enquiry, and we did not capture DVA enquiry in subsequent appointments, when rapport may have been established or a partner may not have attended. Thirdly, routinely collected data do not indicate the reasons why midwives were unable to ask about DVA. While we can speculate on the reasons, further research is needed to explore these and identify pathways for service improvement. Finally, many potential predictors (such as staffing, setting or service-level factors) are not routinely recorded, which may provide insights into the main barriers to DVA enquiry.

In conclusion, despite recommendations for universal enquiry, this study found that individuals from certain groups are more likely to be asked about DVA victimisation than others at antenatal appointments. This highlights opportunities for training and system interventions to optimise identification and response to DVA among all women accessing antenatal care.

## Data Availability

The data underlying this study are not publicly available because they contain sensitive patient-level information and are subject to legal and contractual restrictions imposed by the data provider (GSTT NHS Trust). Although the data have been pseudoanonymised, they include detailed personal and medical variables that could enable re-identification and are deemed to be sensitive. The authors accessed the data under a data sharing agreement for a specific third-party project and are not permitted to share the dataset. Access may be granted to qualified researchers by the NHS Trust, subject to appropriate governance approvals (including ethical approval and data sharing agreements).

## Acknowledgements

We thank the LEAP team and families for enabling this research. We are grateful to the patients whose anonymised health records contributed to this research.

